# The impact of self-isolation due to COVID-19 on health care workers’ mental health and wellbeing: a systematic review with narrative synthesis

**DOI:** 10.1101/2025.02.18.25322220

**Authors:** Madeline V. Stein, Samantha K. Brooks, Louise E. Smith, G. James Rubin, Richard Amlôt, Neil Greeberg, Alex F. Martin

**Author notes:** Correspondence address: Madeline V. Stein Department of Psychology Institute of Psychiatry, Psychology & Neuroscience King’s College London 16 De Crespigny Park London SE5 8AB, UK. For open access purposes, the author will apply a Creative Commons Attribution (CC BY) license to any Author Accepted Manuscript version arising.

## Abstract

**Background:** Self-isolation is a key public health strategy for infectious disease control. Globally implemented during the COVID-19 pandemic, it remains an essential strategy in ongoing mitigation efforts. Healthcare workers (HCWs) often face isolation due to occupational exposure to infectious diseases and may face unique psychological challenges.

**Aims:** This systematic review synthesized evidence on (1) the impact of isolation on HCWs’ psychological wellbeing, (2) factors associated with wellbeing, and (3) the effectiveness of interventions to improve wellbeing during or after isolation for COVID-19.

**Methods:** A pre-registered systematic review (PROSPERO: CRD42024559971) was conducted in accordance with PRISMA and Cochrane guidelines. Searches in PsycInfo, Embase, MEDLINE, PubMed, and grey literature included studies on HCWs’ psychological wellbeing during or after self-isolation. Risk of bias was assessed using ROBINS-E or CASP tools.

**Results:** From 20,798 records screened, 19 studies (10 quantitative, 7 qualitative, 2 mixed methods) were included. Quantitative findings on anxiety, depressive, and stress symptoms were inconsistent. Qualitative studies consistently reported distress, loneliness, and stigma. Factors associated with wellbeing included socio-cultural influences and protective factors. No studies assessed interventions targeting wellbeing during self-isolation.

**Conclusion:** Self-isolation appears to have variable effects on HCWs’ wellbeing, including significant challenges and opportunities for resilience. Public health strategies should prioritize timely, clear communication, accessible evidence-based psychological support, and practical resources. Future research must prioritize evaluation of interventions to mitigate psychological harm and support HCWs during infectious disease outbreaks.

**Teaser text:** Self-isolation is vital for infectious disease control but poses psychological challenges for healthcare workers (HCWs). We synthesized findings on the impact of self-isolation on HCWs’ wellbeing, associated factors, and interventions to enhance wellbeing. Quantitative studies reported inconsistent wellbeing findings, while qualitative data highlighted distress, stigma, and resilience. Public health strategies must ensure psychological and practical support, clear communication, and intervention development to mitigate harm and enhance HCWs’ wellbeing during future infectious disease outbreaks.

## Introduction

During the COVID-19 pandemic, isolation and quarantine were globally implemented public health strategies to control the spread of the virus. Isolation is the separation of those who are sick from those who are well; while quarantine is the separation of those who have been exposed to an illness but are not yet symptomatic. Both measures are critical strategies to curb the transmission of infectious diseases (1). For the purpose of this review the term ‘self-isolation’ encompasses both isolation and quarantine, distinct from broader population-wide ‘lockdowns’. A previous review showed that self-isolation was associated with generally negative mental health and wellbeing outcomes for individuals in the general population, particularly PTSD (2).

Healthcare workers (HCWs), in the present review defined as licensed physical and mental health professionals, hospital staff, and administrators, were particularly vulnerable to infection during the COVID-19 pandemic due to their occupational exposure (3). The nature of their roles often placed them at significantly higher risk of contracting the virus (4). As such, many healthcare settings implemented self-isolation protocols, especially for HCWs directly exposed to infected individuals, in order to mitigate the spread of infection (5). This included post-shift self-isolation requirements either at home or in a work-sanctioned isolation facility (e.g., hospital ‘dorms’ or contracted hotels), regardless of whether the individual had been in direct contact with an infected person.

The combination of occupational exposure and self-isolation protocols introduced challenges for HCWs, compounding the psychological burden they faced during the pandemic (2, 6, 7) arising from exposure to occupational trauma, increased risk of infection, repeated periods of self-isolation, and a heightened fear of transmitting the virus to family members (8–10).

Consequently, HCWs may have experienced distinct psychological outcomes compared to the general population, including moral injury, trauma, and burnout (11, 12). Understanding these unique stressors and outcomes is essential for shaping future public health strategies, particularly as self-isolation remains a key measure in managing infectious diseases (e.g., 2022 mpox outbreak) (8) and for the treatment of diseases that present a significant burden globally, such as tuberculosis (13).

To address these gaps in the literature, we conducted a pre-registered systematic review with three primary aims: (1) to synthesize and evaluate the impact of self-isolation on psychological and emotional wellbeing (hereafter wellbeing) of HCWs during the COVID-19 pandemic; (2) to identify factors associated with wellbeing outcomes in HCWs during or after self-isolation; and (3) to assess the effectiveness of interventions designed to improve the wellbeing of HCWs during or following self-isolation. To achieve these aims, we systematically identified and appraised original studies, narratively synthesizing their findings to provide a comprehensive overview of the current evidence.

## Methods

We completed this prospectively registered review (PROSERO registration number: CRD42024559971) in accordance with Cochrane Collaboration guidelines (14) and the Preferred Reporting Items for Systematic Reviews and Meta-Analyses (15).

### Inclusion criteria

The papers subjected to synthesis must have been published on or after 01 January 2020 and contain original data relating to the psychological wellbeing of HCWs during or after COVID-19-related self-isolation. HCWs may be identified as the main population of interest, or within a broader study where data from HCWs is reported separately from that of the general population. We excluded studies where HCWs were placed into isolation in a hospital or healthcare setting for treatment of COVID-19, rather than as a precaution to prevent the spread of COVID-19. For studies that assessed the impact of self-isolation (Aim 1), they must have employed a pre-exposure comparison (baseline, within-subjects) or control group comparison (non-isolated, between-groups). In studies that assessed factors associated with wellbeing outcomes (Aim 2), we included studies that looked at factors occurring during (e.g., communication quality, individual difference factors, etc.) or related directly (e.g., duration of isolation) to the self-isolation period. In studies that assessed the efficacy of an intervention to improve wellbeing (Aim 3), the study design must have reported pre- and post-intervention scores in the intervention group and/or a control group comparison.

### Search strategy

Medline, PsycInfo, Web of Science, Embase, PsyArXiv, and medRxiv were systematically searched. An initial search was performed in December 2022 by AFM, the search was repeated in August 2023 and May 2024 by MVS, resulting in two new studies for inclusion. The search string (see https://tinyurl.com/ymy363k2) was augmented with manual searches of all included articles’ reference lists and relevant reviews captured in the initial search. We previously searched five grey literature databases for intervention studies only (2) but did not update this search for the present review, as no studies were identified through this method from previous searches.

### Study selection

The initial screening was conducted as part of a complementary review looking at wellbeing in the adult population (2). In the previous review, the search was piloted, and the screening team (AFM, LES, SKB, MVS, and GJR) reviewed a training set of 300 studies. Discrepancies were discussed until agreement on included studies was attained. During the formal study selection process, one team member (AFM) set aside articles that met the current review’s inclusion criteria and confirmed study selection with the screening team. The authors of three potentially eligible papers were contacted for methodological clarification; these papers were excluded because they did not meet the inclusion criteria.

### Data extraction

Data extraction was performed by one reviewer (MVS for quantitative and SKB for qualitative), all extracted data are available open access (see https://osf.io/jngme/). During data extraction, any uncertainties were discussed with the screening team. The data subjected to extraction were defined *a priori* and included: A) study characteristics: i, country; ii, dates of data collection; iii, study design and data collection method; iv, sampling details; v, inclusion criteria and final sample details (age, sex, etc.). B) isolation characteristics: i, reason and location of self-isolation; ii, assessment points; iii, temporal proximity of data collection to self-isolation period. C) quantitative outcomes by research aim: i, outcome measure, ii, analysis type, iii, significant associations and no evidence for significant associations. D) qualitative outcomes by research aim: i, measure; ii, analysis method; iii, impact of isolation; iv, factors perceived to be associated; v, other potentially relevant information.

### Methodological quality assessment

Study quality was assessed by the same two reviewers for each study type. Quantitative study quality was assessed using either the Risk of Bias in Non-randomized Studies for Exposure (ROBINS-E) (16) or Interventions (ROBINS-I) (17). Papers reporting data addressing more than one of our review’s aims received a ROBINS score for each aim. Studies were rated as low risk, some concerns, high risk, or very high risk based on the tool’s algorithm. Qualitative studies were assessed using a modified version of the Critical Appraisal Skills Programme (CASP) checklist, a ten-item measure of study quality (18). The item ‘how valuable is the research?’ was reworded to ‘do the authors discuss the value of the research in terms of implications and contribution to literature?’. This allowed for binary responses (Yes=1, No=0) in line with the other items. An overall quality percent score was computed for each study, with higher scores indicating better quality.

### Study synthesis

Given the research aim of appraising the significance and direction of the relationship between self-isolation and wellbeing, effect sizes were not reported. To capture effects across diverse methodologies we prospectively planned to narratively synthesise the results following guidance from Chapter 12 of the Cochrane Handbook (McKenzie & Brennan, 2023). Quantitative studies were narratively synthesized in accordance with synthesis without meta-analysis (SWiM) guidelines (19). Qualitative studies were synthesized using meta-ethnography following eMERGe guidelines (20). Syntheses were partitioned *a priori* by review aim and grouped by psychological outcome, further grouping categories were defined post-hoc as part of the process of synthesis.

## Results

A PRISMA diagram presenting study selection can be found in Fig. 1. The final sample included 19 papers, the details of which are summarized in Table 1. A full list of included papers is available in the open access data file (OSF link). A total of eighteen studies reported findings related to the impact of self-isolation on the wellbeing of healthcare workers (Aim 1). These included nine quantitative studies, seven qualitative studies, and the qualitative components of two mixed-methods studies. Fourteen studies reported factors associated with wellbeing during or after self-isolation (Aim 2). These included three quantitative studies, one of which addressed this aim exclusively, as well as the quantitative components of two mixed-methods studies. Additionally, five qualitative studies and the qualitative components of two mixed-methods studies addressed Aim 2. No studies investigated the effectiveness of interventions aimed at improving the wellbeing of healthcare workers during or after self-isolation (Aim 3).

**Figure 1.**
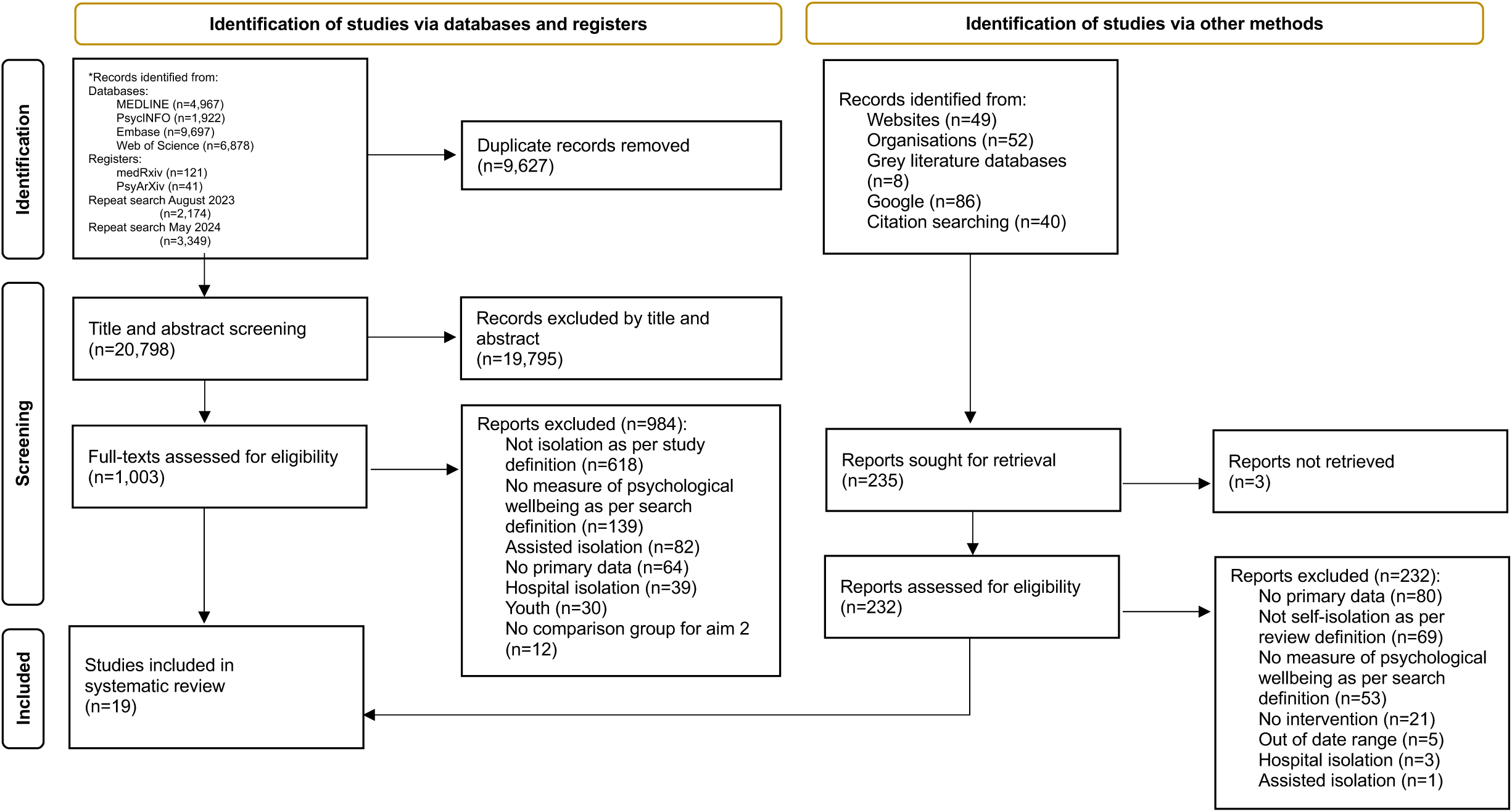
PRISMA flowchart of the study selection process.

**Table 1.**
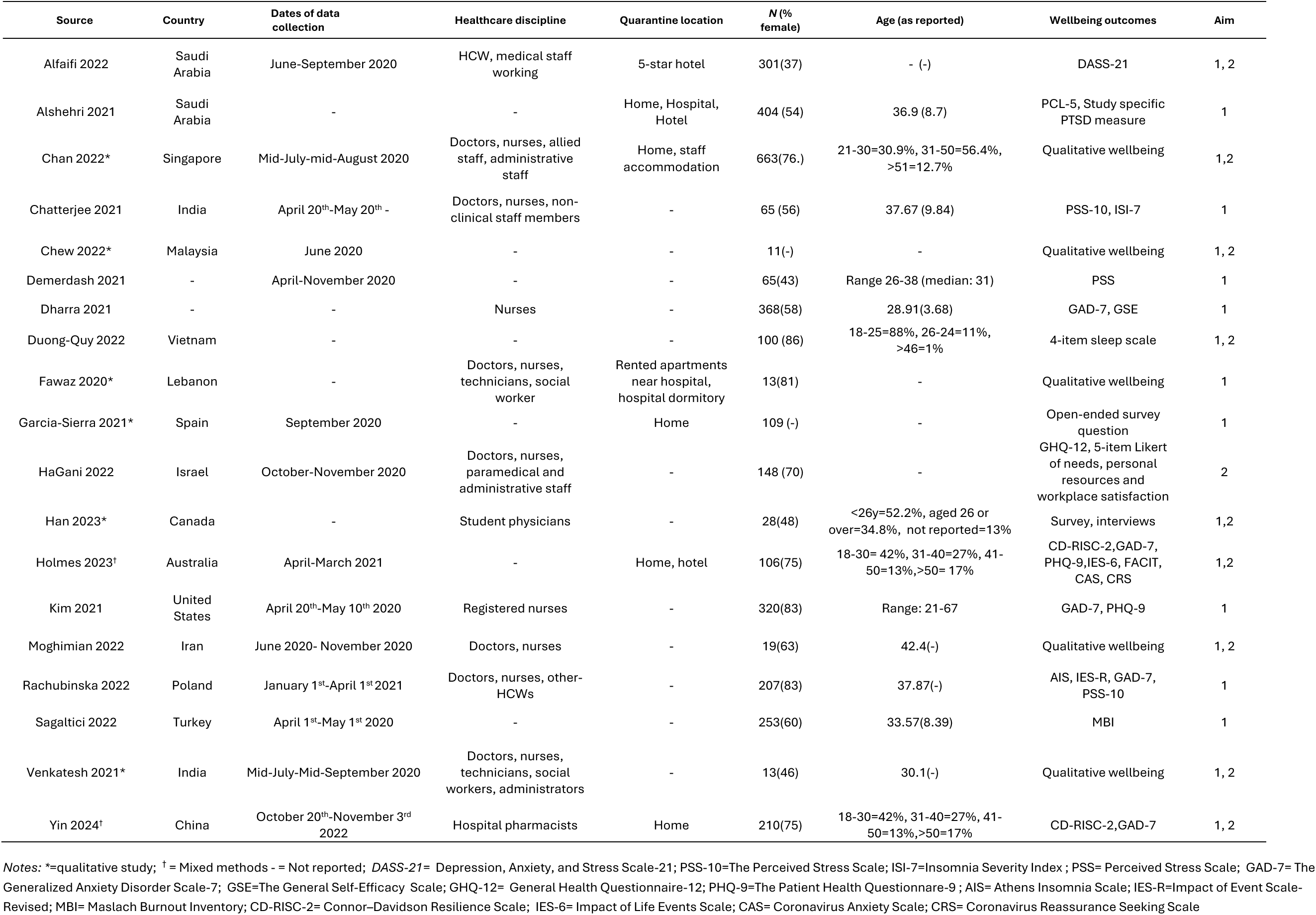
Principal features of included quantitative and qualitative papers.

### The impact of self-isolation on wellbeing (Aim 1)

The quantitative impact of self-isolation on wellbeing was reported in nine studies. Two studies were ‘very high risk of bias’ and the rest were ‘high risk’ (Figure 2). Evidence for any impact of self-isolation on wellbeing was variable. Three studies reported a significant association between anxiety symptoms and self-isolation (21–23), while two studies found no significant association (24, 25). Similarly, evidence regarding depression was inconsistent: two studies identified a significant association with self-isolation (21, 25), whereas one study did not (23).

**Figure 2.**
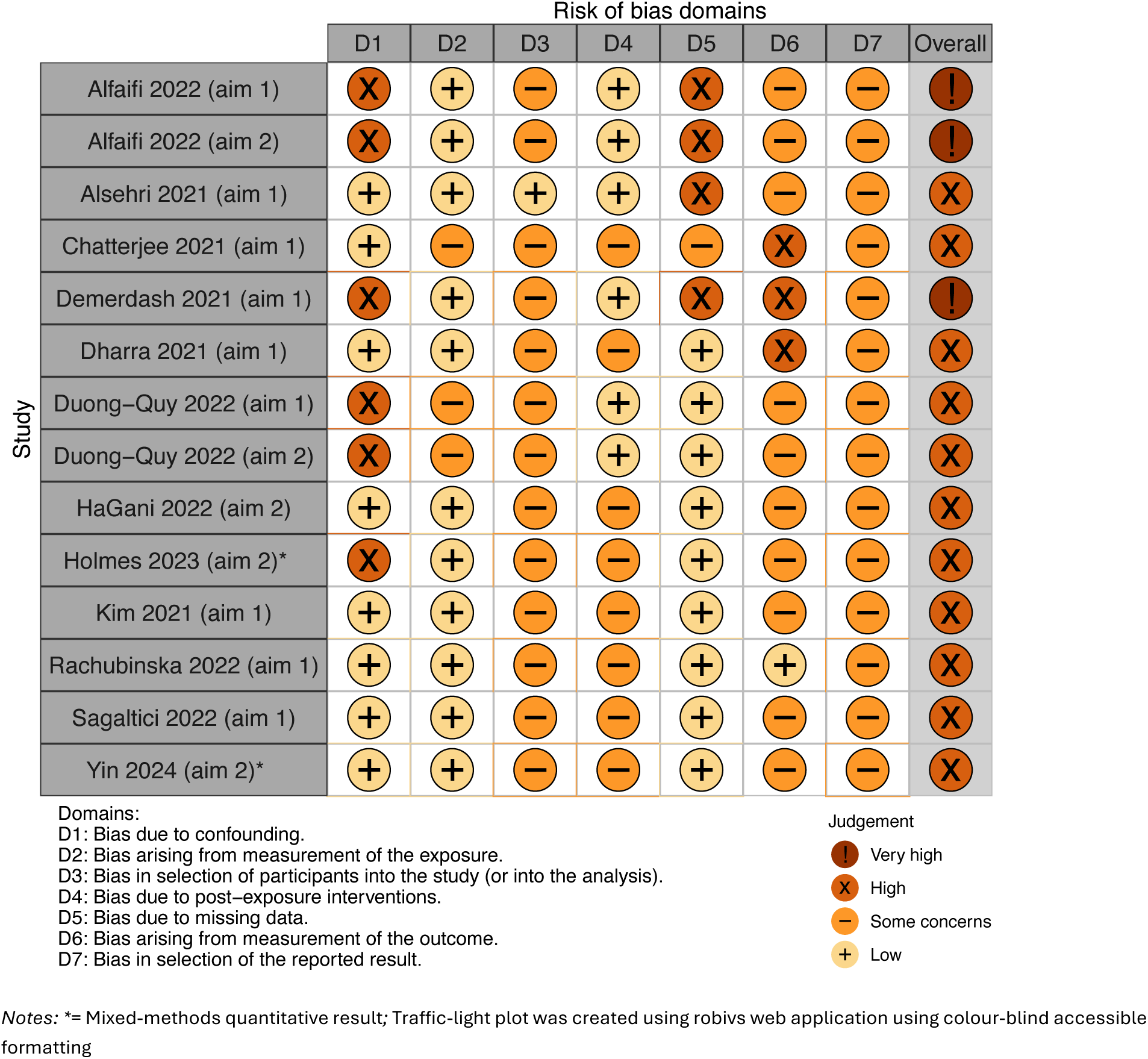
Summary of risk of bias in quantitative studies assessed with the ROBINS-E.

Stress was assessed in three studies: one study found that stress levels were higher during self-isolation (26) while two studies found no association (23, 24). Quarantine was found to improve sleep quality in one study (27) and decrease it in another (24). Other psychological outcomes, such as burnout (28), hopelessness (24), and PTSD symptoms (29) were assessed in a single study and thus were unable to be synthesized.

Nine qualitative studies investigated the impact of self-isolation on wellbeing. Overall, the methodological quality of qualitative studies was satisfactory, on average all studies met over half of the assessed quality criteria (M ± SD: 73% ± 10%; Range: 60–90%). The qualitative findings revealed a wide range of negative psychological and emotional effects experienced by participants during self-isolation. Emotional distress was often reported, with individuals describing feelings of loneliness, sadness, anxiety, tension, and general distress (30–35). Participants expressed fears related to the pandemic, including fears of contracting COVID-19 or potentially spreading it to others, fears of death, and fears of being judged or stigmatised (31, 32, 36–38). Worries about the health of family and friends, along with financial concerns, were also significant sources of stress (31, 33, 36).

The qualitative findings also identified positive effects of self-isolation. Some participants described having time for rest and relaxation (33, 36), a renewed sense of gratitude, and an appreciation for small joys in life, with some feeling they had been given a second chance (32, 36). Others highlighted quick adaptation to the situation (34) and newfound perspectives on their professional roles, including increased empathy for patients and a stronger sense of professional commitment (31, 36).

### Factors associated with wellbeing (Aim 2)

Factors associated with wellbeing were reported in four quantitative and seven qualitative studies. There was substantial heterogeneity in reported outcomes across methodologies assessing factors associated with wellbeing. Factors associated with wellbeing spanned a broad range of socio-cultural and psychological domains, including risk factors for adverse wellbeing outcomes and factors that protect wellbeing. One quantitative finding was ‘very high risk of bias’ and the rest were ‘high risk’ (Figure 2). As with Aim 1, qualitative findings were of satisfactory quality (M ± SD: 75% ± 10%; Range: 60–90%).

### Risk factors

In the quantitative studies, greater COVID-19 stigma was associated with worse wellbeing outcomes (21, 39). Similarly, longer quarantine duration was associated with worse wellbeing outcomes in two studies (37, 39), while one study did not find a significant relationship (21). Finally, bed quality was associated with sleep improvement in one study (27).

In the qualitative studies, participants discussed specific situations and characteristics which they believed made self-isolation more difficult. Having recently moved and still adapting to new circumstances was reported to make self-isolation harder (30), while experiencing other emergencies during the self-isolation period (e.g. being unable to take an unwell child to the doctor) negatively affected mental state (34). Receiving conflicting information contributed to uncertainty (31). The perception of being stigmatized by others made the situation more distressing (30). Perceiving the mitigation measures to be too strict also appeared to negatively affect wellbeing (32). Inadequate basic supplies in quarantine and/or unmet medical needs also made the situation more difficult to cope with (34, 37). Other challenges reported by Holmes, Ellen (37) included lack of physical exercise; financial impact of quarantine; and feeling pressure to continue working. In addition to these emotional and psychological impacts, participants noted experiencing sleep disturbances, including nightmares, which were linked to heightened anxiety (36). Some individuals highlighted the challenges of boredom and loneliness, which were pervasive during self-isolation and compounded feelings of isolation and disconnection (31, 33, 36, 37). Furthermore, a sense of uncertainty about the future and their circumstances added to the psychological burden (31).

### Protective factors

One quantitative study found resilience mediated a decrease in anxiety (34), whereas another study found no association between resilience and wellbeing (37). Additionally, positive opinions on pandemic prevention were found to be associated with resilience in one study (37). Qualitative participants described coping strategies which were perceived to make self-isolation easier, including religious practices (33); support from others (31, 32, 34, 36); exercise (34); keeping a positive mindset (34); doing activities relating to work while in self-isolation, such as reading relevant literature (32); and self-care activities (34, 36).

## Discussion

The present review summarized and appraised the available literature assessing the impact of self-isolation on HCWs psychological wellbeing during the COVID-19 pandemic. Overall, the included studies represent a range of isolation contexts, healthcare disciplines, and countries. Quantitative outcomes and evidence were heterogenous, and of ‘high’ or ‘very high risk of bias’, with an inconsistent trend towards an increase in anxiety, depressive, and stress symptoms. The qualitative studies were of satisfactory methodological quality and indicated that while multiple factors negatively affected HCWs’ wellbeing during isolation, there were also factors that improved and protected wellbeing. Finally, no studies assessed the efficacy of interventions to increase HCWs’ wellbeing during isolation, highlighting an important area of research during future infectious disease outbreaks.

Across methodologies, our results cumulatively suggest that self-isolation had a negative impact on HCWs’ wellbeing during the COVID-19 pandemic. Quantitative studies primarily focused on psychopathology using self-report measures, with most studies assessing anxiety, depressive, and stress symptoms. However, these scales often fail to capture broader aspects of wellbeing, such as frustration and agitation, which are critical for understanding factors such as government mistrust and reduced adherence to mitigation strategies (40, 41). Furthermore, although it was rarely reported and not formally extracted, HCWs’ isolation may have been employer-mandated, introducing unique workplace-related dynamics that could influence satisfaction and fulfilment.

Indeed, we anticipated that occupational exposures and isolation would likely result in adverse outcomes such as moral injury in HCWs, but none of the studies included in our review assessed this. While qualitative evidence showed that many participants reported negative psychological effects, most prominently fear (e.g., of the disease or potentially spreading it) and loneliness, several studies also reported evidence of potential post-traumatic growth (e.g., renewed sense of gratitude and new perspective on patient care). As such, despite reports of several factors which were perceived to make the situation more challenging (e.g., stigma, financial stress, etc.) there were also factors perceived to make self-isolation easier (e.g., support from others).

These findings align with those reported in a previous review of the general population (2). In the present review and that of the general population, qualitative studies provided a more nuanced understanding of self-isolation’s impact on wellbeing, capturing broad domains such as fear (present review) and worry (general population review) (2). Protective factors were identified in both reviews, with social support and coping strategies serving as key buffers for both HCWs and the general population. However, a notable discrepancy emerged between the current review and that of the general population: while PTSD symptoms were a consistent concern in the general population, our review could not synthesize PTSD symptoms as a psychological outcome due to the limited number of available studies (n=1). Additionally, protective factors were more comprehensively explored in our review compared to that of the general population. This discrepancy is plausibly due to the availability of primary research within each review and highlights the need for further research to better characterize PTSD-specific risk to HCWs, ensuring a more comprehensive understanding of the psychological impact of self-isolation.

### Limitations and future directions

The results of this review should be interpreted in light of the strengths and limitations of both the included articles and the review process itself.

All the quantitative studies were rated as ‘high’ or ‘very high risk of bias’. Most studies (N=12) showed at least ‘some concerns’ regarding the selection of sample participants, and all studies were rated as having at least ‘some concerns’ related to bias in their selected results. This was typically due to a lack of pre-registration. During future infectious disease outbreaks, researchers should prioritize pre-registering their study protocols and planned analyses to enhance transparency and reliability. Similarly, three of the quantitative findings relied on study-specific measures (e.g., 4-item Likert sleep scale, 5-item Likert assessing needs, personal resources, and workplace satisfaction, and unvalidated PTSD measure) which limited the reliability and validity of their results. Future studies should prioritize the use of established, validated measures. Finally, the quantitative results were mixed, with no outcome domain reaching consensus. This variability may be attributable to study-specific contexts. For instance, one study assessing sleep quality found that isolation improved sleep (27), likely reflecting the sample’s specific characteristics, where pre-isolation working conditions had adversely affected sleep duration and quality. Future studies should consider and clearly report contextual factors, such as pre-isolation conditions, to improve comparability and generalizability of findings across diverse populations.

Qualitative studies were of satisfactory methodological quality overall, but there were several items on the quality appraisal checklist where studies most often lost points. First, none of the studies discussed reflexivity, or considered how the researchers’ own expectations, perceptions or biases may have influenced data collection or analysis. Second, in at least four studies it was unclear whether data analysis was sufficiently rigorous, and several studies did not describe their analytic procedure in enough detail for the study to be replicable. Finally, it was unclear for most studies whether recruitment strategies were appropriate. Future research should aim to address these limitations and ensure they are clearly discussed within the text.

No studies in this review assessed the efficacy of interventions, psychological or practical, aimed at improving HCWs’ wellbeing during isolation. This represents a significant gap in the literature, as targeted interventions will play a crucial role in mitigating the psychological and emotional challenges faced by HCWs during future infectious disease outbreaks. Addressing this gap through rigorous research is essential to inform evidence-based strategies that enhance HCWs’ resilience and wellbeing during future infectious disease outbreaks.

Our review process had many strengths, such as a pre-registered protocol, no geographic or language limitations, comprehensive risk of bias assessment and adherence to PRISMA, SWiM, and eMERGe guidelines. However, human error may have resulted in missed studies during the selection process. Similarly, while data extraction was discussed with the review team, the absence of a dual-coder process may have increased the risk of errors.

### Conclusions

This review provides a summary of the current evidence on the impact of self-isolation on healthcare workers’ psychological wellbeing during the COVID-19 pandemic. While qualitative findings highlighted both challenges and growth opportunities during self-isolation, quantitative studies presented inconsistent evidence, trending toward negative psychological effects such as increased anxiety, depressive, and stress symptoms. The absence of research on interventions to improve HCWs’ wellbeing during self-isolation represents a critical gap that should be addressed before future infectious disease outbreaks. Public health strategies should prioritize timely, clear communication, accessible psychological and practical support, and resources to mitigate psychological harm. Future research should address methodological limitations in the current evidence base by incorporating pre-registration, validated measures, and workplace-specific stressors and outcomes to enhance reliability and relevance. By closing these gaps, research can provide actionable insights to better support HCWs and enhance their psychological wellbeing during public health crises.

## Author contributions

AFM, LES, RA, NG and GJR conceptualized the study. AFM ran the systematic searches and MVS updated the searches. MVS, SKB, LES, GJR, and AFM screened citations. MVS and SKB completed data extraction and risk of bias ratings with support from all authors. MVS drafted the initial manuscript with support from AFM and SKB. All authors reviewed and edited the manuscript prior to submission.

## Data availability statement

The data are freely available in all included articles. Extracted data can be freely accessed: https://osf.io/jngme/

## Supporting information

see https://tinyurl.com/ymy363k2

## References

1. Wilder-Smith A, Freedman DO. Isolation, quarantine, social distancing and community containment: pivotal role for old-style public health measures in the novel coronavirus (2019-nCoV) outbreak. Journal of travel medicine. 2020;27(2):taaa020.

2. Martin AF, Smith LE, Brooks SK, Stein MV, Davies R, Amlôt R, et al. The impact of self-isolation on psychological wellbeing and how to reduce it: a systematic review. medRxiv. 2023:2023.10.16.23296895.

3. Chou R, Dana T, Buckley DI, Selph S, Fu R, Totten AM. Epidemiology of and Risk Factors for Coronavirus Infection in Health Care Workers. Annals of Internal Medicine. 2020;173(2):120–36.

4. Dzinamarira T, Nkambule SJ, Hlongwa M, Mhango M, Iradukunda PG, Chitungo I, et al. Risk Factors for COVID-19 Infection Among Healthcare Workers. A First Report From a Living Systematic Review and meta-Analysis. Safety and Health at Work. 2022;13(3):263–8.

5. UK Health Security Agency. 2020. [16 November 2022]. Available from: https://ukhsa.blog.gov.uk/2020/02/20/what-is-self-isolation-and-why-is-it-important/.

6. Brooks SK, Webster RK, Smith LE, Woodland L, Wessely S, Greenberg N, et al. The psychological impact of quarantine and how to reduce it: rapid review of the evidence. The lancet. 2020;395(10227):912–20.

7. Gómez-Durán EL, Martin-Fumadó C, Forero CG. Psychological impact of quarantine on healthcare workers. Occup Environ Med. 2020;77(10):666–74.

8. Saragih ID, Tonapa SI, Saragih IS, Advani S, Batubara SO, Suarilah I, et al. Global prevalence of mental health problems among healthcare workers during the Covid-19 pandemic: A systematic review and meta-analysis. Int J Nurs Stud. 2021;121:104002.

9. Greene T, Harju-Seppanen J, Billings J, Brewin CR, Murphy D, Bloomfield MAP. Exposure to potentially morally injurious events in U.K. health and social care workers during COVID-19: Associations with PTSD and complex PTSD. Psychol Trauma. 2023.

10. Kapetanos K, Mazeri S, Constantinou D, Vavlitou A, Karaiskakis M, Kourouzidou D, et al. Exploring the factors associated with the mental health of frontline healthcare workers during the COVID-19 pandemic in Cyprus. PLoS One. 2021;16(10):e0258475.

11. Kwaghe AV, Ilesanmi OS, Amede PO, Okediran JO, Utulu R, Balogun MS. Stigmatization, psychological and emotional trauma among frontline health care workers treated for COVID-19 in Lagos State, Nigeria: a qualitative study. BMC Health Services Research. 2021;21(1):855.

12. Bielicki JA, Duval X, Gobat N, Goossens H, Koopmans M, Tacconelli E, et al. Monitoring approaches for health-care workers during the COVID-19 pandemic. Lancet Infect Dis. 2020;20(10):e261–e7.

13. Falzon D, Miller C, Law I, Floyd K, Arinaminpathy N, Zignol M, et al. Managing tuberculosis before the onset of symptoms. Lancet Respir Med. 2025;13(1):14–5.

14. Higgins JPT, Higgins JPT, Green S, Cochrane C. Cochrane handbook for systematic reviews of interventions. Cochrane book series. Chichester Oxford: John Wiley & Sons Ltd Cochrane Collaboration; 2008.

15. Page MJ, Moher D, Bossuyt PM, Boutron I, Hoffmann TC, Mulrow CD, et al. PRISMA 2020 explanation and elaboration: updated guidance and exemplars for reporting systematic reviews. BMJ. 2021;372:n160.

16. Higgins J, Morgan R, Rooney A, Taylor K, Thayer K, Silva R, et al. Risk Of Bias In Non-randomized Studies-of Exposure (ROBINS-E): Launch version. Risk Of Bias. 2022.

17. Sterne JA, Hernán MA, Reeves BC, Savović J, Berkman ND, Viswanathan M, et al. ROBINS-I: a tool for assessing risk of bias in non-randomised studies of interventions. bmj. 2016;355.

18. CASP Qualitative Checklist [Internet]. 2022 [cited 12 April 2023]. Available from: https://casp-uk.net/casp-tools-checklists/.

19. Campbell M, McKenzie JE, Sowden A, Katikireddi SV, Brennan SE, Ellis S, et al. Synthesis without meta-analysis (SWiM) in systematic reviews: reporting guideline. bmj. 2020;368.

20. France EF, Cunningham M, Ring N, Uny I, Duncan EA, Jepson RG, et al. Improving reporting of meta-ethnography: the eMERGe reporting guidance. BMC medical research methodology. 2019;19:1–13.

21. Alfaifi A, Darraj A, El-Setouhy M. The Psychological Impact of Quarantine During the COVID-19 Pandemic on Quarantined Non-Healthcare Workers, Quarantined Healthcare Workers, and Medical Staff at the Quarantine Facility in Saudi Arabia. Psychol Res Behav Manag. 2022;15:1259–70.

22. Dharra S, Kumar R. Promoting Mental Health of Nurses During the Coronavirus Pandemic: Will the Rapid Deployment of Nurses’ Training Programs During COVID-19 Improve Self-Efficacy and Reduce Anxiety? Cureus. 2021;13(5):e15213.

23. Rachubińska K, Cybulska AM, Sołek-Pastuszka J, Panczyk M, Stanisławska M, Ustianowski P, et al. Assessment of Psychosocial Functioning of Polish Nurses during COVID-19 Pandemic. Int J Environ Res Public Health. 2022;19(3).

24. Chatterjee SS, Chakrabarty M, Banerjee D, Grover S, Chatterjee SS, Dan U. Stress, Sleep and Psychological Impact in Healthcare Workers During the Early Phase of COVID-19 in India: A Factor Analysis. Front Psychol. 2021;12:611314.

25. Kim SC, Quiban C, Sloan C, Montejano A. Predictors of poor mental health among nurses during COVID-19 pandemic. Nurs Open. 2021;8(2):900–7.

26. Demerdash HM, Omar E, Arida E. Evaluation of copeptin and psychological stress among healthcare providers during COVID-19 pandemic. Egyptian Journal of Anaesthesia. 2021;37(1):227–33.

27. Duong-Quy S, Tran-Duc S, Hoang-Chau-Bao D, Bui-Diem K, Vu-Tran-Thien Q, Nguyen-Nhu V. Tiredness, depression, and sleep disorders in frontline healthcare workers during COVID-19 pandemic in Vietnam: A field hospital study. Front Psychiatry. 2022;13:984658.

28. Sagaltici E, Saydam RB, Cetinkaya M, Şahin Ş K, Küçük SH, Müslümanoğlu AY. Burnout and psychological symptoms in healthcare workers during the COVID-19 pandemic: Comparisons of different medical professions in a regional hospital in Turkey. Work. 2022;72(3):1077–85.

29. Alshehri AS, Alghamdi AH. Post-traumatic Stress Disorder Among Healthcare Workers Diagnosed With COVID-19 in Jeddah, Kingdom of Saudi Arabia, 2020 to 2021. Cureus. 2021;13(8):e17371.

30. Chan AY, Ting C, Chan LG, Hildon ZJ-L. “The emotions were like a roller-coaster”: a qualitative analysis of e-diary data on healthcare worker resilience and adaptation during the COVID-19 outbreak in Singapore. Human Resources for Health. 2022;20(1):60.

31. Han S, Kim I, Rojas D, Nyhof-Young J. Investigating the experiences of medical students quarantined due to COVID-19 exposure. Can Med Educ J. 2023;14(6):92–101.

32. Moghimian M, Farzi K, Farzi S, Moladoost A, Safiri S. Exploring the experiences of nurses and physicians infected with COVID-19. J Educ Health Promot. 2022;11:35.

33. Chew KS, Ibrahim N, Fazillah NAM, Subramaniam D, Sibey AV, Silaiman FN, et al. Lived experiences and coping responses toward mandatory quarantine among Malaysian healthcare workers during COVID-19 pandemic: A qualitative analysis. Geografia. 2022;18(2):200–9.

34. Yin Z, Wang X, Lu X, Fu H. Hospital pharmacists’ mental health during home isolation in the post-pandemic era of COVID-19: influencing factors, coping strategies, and the mediating effect of resilience. Front Public Health. 2024;12:1268638.

35. García-Sierra R, Moreno-Gabriel E, Badia Perich E, Sabaté Cintas V, Bonet Simó JM, Violán CF, et al. Evaluation of a support system for health professionals confined by COVID-19. Rev Saude Publica. 2021;55:108.

36. Venkatesh V, Samyuktha VN, Wilson BP, Kattula D, Ravan JR. Psychological impact of infection with SARS-CoV-2 on health care providers: A qualitative study. J Family Med Prim Care. 2021;10(4):1666–72.

37. Holmes OS, Ellen S, Smallwood N, Willis K, Delaney C, Worth LJ, et al. The Psychological and Wellbeing Impacts of Quarantine on Frontline Workers during COVID-19 and Beyond. Int J Environ Res Public Health. 2023;20(10).

38. Fawaz M, Samaha A. The psychosocial effects of being quarantined following exposure to COVID-19: A qualitative study of Lebanese health care workers. Int J Soc Psychiatry. 2020;66(6):560–5.

39. HaGani N, Eilon Y, Zeevi S, Vaknin L, Baruch H. The psychosocial impact of quarantine due to exposure to COVID-19 among healthcare workers in Israel. Health Promot Int. 2023;38(3).

40. Choi KW, Jung JH, Kim HH-s. Political trust, mental health, and the coronavirus pandemic: A cross-national study. Research on Aging. 2023;45(2):133–48.

41. Smith LE, Amlȏt R, Lambert H, Oliver I, Robin C, Yardley L, et al. Factors associated with adherence to self-isolation and lockdown measures in the UK: a cross-sectional survey. Public Health. 2020;187:41–52.

