## Supplementary material for "The impact of self-isolation due to COVID-19 on health care workers’ mental health and wellbeing: a systematic review with narrative synthesis": see https://tinyurl.com/ymy363k2

| | |
|---|---|
| # | **Query** |
| 1 | (coronavirus or covid* or sars-cov-2 or ncov2019).mp. or exp Coronavirus/ or exp COVID-19/ or exp SARS-CoV-2/ |
| 2 | (isolat* or quarantin* or confinement).mp. or exp Patient Isolation/ or exp Quarantine/ |
| 3 | 2 not "social isolation".mp. [mp=title, book title, abstract, original title, name of substance word, subject heading word, floating sub-heading word, keyword heading word, organism supplementary concept word, protocol supplementary concept word, rare disease supplementary concept word, unique identifier, synonyms] |
| 4 | (adheren* or compliance or wellbeing or well-being or "quality of life" or resilien* or coping or flourish* or "positive psychology" or "posttraumatic growth" or "post-traumatic growth" or "life satisfaction" or "personal satisfaction" or "psychosocial functioning" or "mental health" or anxiety or depress* or ptsd or trauma* or psychiatric or "psychological stress" or "social stigma" or distress* or mood* or emotion* or "substance abuse" or "substance misuse" or "substance use" or "hazardous drinking" or "alcohol use" or "alcohol abuse" or "alcohol misuse" or alcoholi* or sleep or insomnia or loneliness).mp. or exp Guideline Adherence/ or exp "Treatment Adherence and Compliance"/ or exp Compliance/ or exp Patient Compliance/ or exp "Quality of Life"/ or exp Resilience, Psychological/ or exp Psychology, Positive/ or exp Posttraumatic Growth, Psychological/ or exp Personal Satisfaction/ or exp Psychosocial Functioning/ or exp Mental Health/ or exp Anxiety Disorders/ or exp Anxiety/ or exp Panic/ or exp Panic Disorder/ or exp Depression/ or exp Stress Disorders, Post-Traumatic/ or exp Psychological Trauma/ or exp Stress, Psychological/ or exp Social Stigma/ or Psychological Distress/ or exp Emotions/ or exp Sleep/ or exp "Sleep Initiation and Maintenance Disorders"/ or exp Substance Abuse, Intravenous/ or exp Substance-Related Disorders/ or exp Alcoholism/ or exp Alcohol Drinking/ or exp Loneliness/ |
| 5 | 1 and 3 and 4 |
| 6 | limit 5 to (humans and yr="2020 -Current") |
